# Cost Homogenization and System-Level Drivers in Plateau Laparoscopic Cholecystectomy: Failure and Reconstruction of Traditional Cost-Control Models in the DRG Era

**DOI:** 10.64898/2026.08.17.26360536

**Authors:** Zhongfeng Dang, Guoliang Ren, Zhiqiang Wang, Wei Su, Yabing Ma, Ping Li, Dongde Ji, Liansheng Li, Junlin Gao

**Author notes:** Primary corresponding author: Zhongfeng DANG. Co-corresponding authors: Junlin GAO. Co-first authors (Zhongfeng Dang, Guoliang Ren, Zhiqiang Wang). Co-second authors (Wei Su, Yabing Ma).

## Abstract

**Background:** Under the DRG/DIP payment reform, the cost structure and its driving factors for laparoscopic cholecystectomy (LC) in resource-limited plateau regions remain unclear. Traditional cost-control models focus on clinical process factors such as length of stay and operative time, but their applicability in the DRG era requires validation.

**Methods:** Based on a single-center cohort of 605 plateau LC patients from May 2020 to October 2025, natural log transformation was applied to total hospitalization costs. Pearson/Spearman correlation analysis, multivariate linear regression (two nested strategies: traditional clinical model and system-driven model incorporating year dummies), and quantile regression were used to identify cost drivers. VIF testing was performed for multicollinearity, and scenario simulation was conducted based on the regression model (operative time reduction to 40 min set according to best historical surgeon efficiency at our center).

**Results:** The mean hospitalization cost was ¥8097.49±936.85 with a coefficient of variation of only 11.6% and a Gini coefficient of 0.062, demonstrating high homogenization. The traditional six-variable clinical model (length of stay, operative time, age, BMI, systolic and diastolic blood pressure) yielded R²=0.008 (F=0.78, P=0.587), with no significant predictors, indicating that traditional clinical factors had largely lost explanatory power for cost variation under DRG control. The system-driven model (incorporating year dummies, season, and clinical variables) achieved R²=0.143 (F=12.41, P<0.001), with year effects (2024: +7.3%, P<0.001; 2025: −5.4%, P=0.001, comprising diagnosis composition changes and DRG policy transition) and seasonal effects (spring: +5.9%, P<0.001) as significant drivers, operative time marginally significant (P=0.039), and length of stay non-significant (P=0.638). Quantile regression revealed that age had a significant positive effect on high-quantile costs (Q90: β=0.0021, P=0.003) but not on median costs. Scenario simulation showed that clinical process optimization (reducing operative time to 40 min) could only reduce costs by approximately 0.6%, far below the annual policy fluctuation (12.1% decline from 2024 to 2025).

**Conclusion:** Plateau LC costs demonstrate high homogenization under DRG control, and traditional clinical process-based cost-control models have minimal explanatory power. System-level factors (annual effects comprising diagnosis composition and policy transition, seasonal resource fluctuations) explain significantly higher cost variation than clinical process factors. We recommend extending the classic Donabedian Structure-Process-Outcome (SPO) framework to a System-Allocation-Outcome (SAO) paradigm, shifting the focus of cost control from clinical process optimization to policy window management and system resource allocation.

## 1. Introduction

Against the backdrop of the comprehensive advancement of Diagnosis-Related Groups (DRG) / Diagnosis-Intervention Packet (DIP) payment reform[1], refined cost control for laparoscopic cholecystectomy (LC) is a core issue in hospital health management[2]. Traditional cost-control studies have predominantly focused on clinical process factors such as length of stay and operative time, and cost-driving models built on non-plateau data have shown that these factors have significant explanatory power for cost variation[2,3]. However, when the institutional environment undergoes a transition (from fee-for-service to DRG standardized pricing), the applicability boundaries of original models may be breached: if costs have already been compressed by standardized grouping at the pricing end, the leverage effect of clinical processes on costs may no longer be significant—that is, the same driving model may exhibit order-of-magnitude differences in explanatory power under different payment systems. DRG payment reform, through standardized grouping and pricing mechanisms, may fundamentally reshape the cost structure and cause fundamental changes in the explanatory power of traditional clinical factors[3]. Plateau regions, characterized by scarce medical resources, high marginal costs of diagnosis and treatment, and significant environment-mediated clinical heterogeneity, may have cost-driving mechanisms that systematically differ from plain regions[4,5]. A plateau cohort of congenital heart disease surgery has reported significant altitude-gradient differences in hospitalization costs[5], and a recent single-center study confirmed that cost-effectiveness analysis of plateau LC has scenario specificity[6], suggesting that plateau surgical cost research requires independent local empirical evidence and should not directly extrapolate plain conclusions. Further empirical research is needed on the characteristics of medical costs in plateau regions.

The classic Donabedian Structure-Process-Outcome (SPO) healthcare quality framework[7] defines patient baseline as structural variables, clinical diagnostic and treatment behaviors as process variables, and outcomes as outcome variables, and has been widely applied in healthcare quality evaluation. However, the applicability of this framework in the DRG era faces challenges: when costs are standardized by DRG pricing, do process variables still have cost leverage effects? Do system-level policy and seasonal factors replace clinical processes as the main cost drivers? The explanation of the same phenomenon often depends on the analytical scale adopted—the micro clinical process perspective and the macro system policy perspective may capture completely different driving signals—this study attempts to test the sources of cost variation explanation at both scales simultaneously through a nested model design. These questions still lack empirical evidence in plateau resource-limited scenarios[3].

This study, based on a cohort of 605 plateau LC patients from May 2020 to October 2025 at Qinghai Red Cross Hospital, systematically tests the explanatory power of traditional clinical factors for costs, identifies the true driving factors of cost variation in the DRG era, and explores the theoretical extension of the SPO framework under DRG cost-control scenarios. The study design follows: variation that cannot be explained by the traditional process model is then examined at the system and resource allocation dimensions, without presetting the hierarchical attribution of driving sources. This study does not preset that traditional clinical process factors still dominate cost variation in the plateau DRG scenario, but treats this as a competing hypothesis to be tested: if the DRG standardized pricing mechanism, by compressing cost variation, causes the explanatory power of clinical process variables to drop by an order of magnitude, then system-level and resource allocation-level signals should be detectable in the nested model (significant R² improvement); conversely, if the R² improvement of the system-driven model is not significant, the traditional hypothesis is retained in this scenario. This competing testing framework ensures that this study can produce definitive scientific conclusions regardless of the direction of results. This study, along with Series I (surgical efficiency analysis) and Series II (epidemiological characteristics analysis), is based on the same clinical cohort but focuses on independent health economic scientific questions (differences in cohort time windows are detailed in Section 2.1), with no risk of duplicate publication.

## 2. Methods

### 2.1 Study Population

Patients undergoing elective LC at our hospital from May 2020 to October 2025 were selected. After excluding records with date anomalies and non-cholelithiasis diagnoses, a final total of 605 patients were included. This study, along with Series I (surgical efficiency analysis, n=591, focusing on the top 7 surgeons by operative volume) and Series II (epidemiological characteristics analysis, n=605), is based on the same clinical cohort; sample size differences arise from distinct analytical objectives. Time window difference explanation: Series I and Series II focus on baseline clinical and efficiency analysis from January 2020 to December 2023; this study, in order to capture the policy transition effects after the comprehensive implementation of DRG in 2024 (annual peak and 2025 cost-control decline), includes consecutively enrolled cases from 2024-2025 (303 cases in the 2020-2023 baseline period are identical to Series I and II, and the 302 newly added cases from 2024-2025 are all from the same center with the same inclusion criteria), ensuring cohort homogeneity. This study is a retrospective anonymized data study. Ethics approval and consent to participate: This study was approved by the Ethics Committee of Qinghai Red Cross Hospital (Approval No.: Lw-2026-73). Written informed consent was waived, and the study strictly follows the ethical principles of the Declaration of Helsinki[8].

### 2.2 Variables

The outcome variable was total hospitalization cost (CNY), which was natural log-transformed before inclusion in regression analysis due to its skewed distribution. Independent variables included: year (2020-2025), season (spring March-May, summer June-August, autumn September-November, winter December-February), length of stay (days), operative time (min), age (years), gender, BMI (kg/m²), systolic blood pressure (mmHg), diastolic blood pressure (mmHg), and preoperative diagnosis classification (simple cholelithiasis vs. cholelithiasis with cholecystitis). Year was included as dummy variables (with 2020-2023 as the reference baseline period, 2024 and 2025 as separate dummy variables), and season was included as a spring dummy variable.

### 2.3 Statistical Analysis

Data analysis was performed using Python 3.11.7 (pandas 2.1.4, statsmodels 0.14.1, scipy 1.11.4). Sample size estimation was based on G*Power 3.1.9.7: with effect size f²=0.15 (medium effect), significance level α=0.05, statistical power 1-β=0.90, and preset number of independent variables k=8, the minimum required sample size was calculated as n=160. The actual sample of 605 patients in this study far exceeds the minimum sample size requirement.

Continuous variables were described as mean±standard deviation, and categorical variables as frequency (percentage). Cost distribution characteristics were assessed using coefficient of variation (CV), interquartile range (IQR), and Gini coefficient. Pearson and Spearman correlation coefficients were used for correlation analysis. Multivariate linear regression employed two nested model strategies: (1) traditional clinical model (length of stay, operative time, age, BMI, systolic blood pressure, diastolic blood pressure—6 variables); (2) system-driven model (adding year dummies [2024, 2025, with 2020-2023 as reference], season, and diagnosis classification to the traditional model variables—8 variables total), with year effects estimated through dummy variables in the system-driven model. Quantile regression (Q10, Q25, Q50, Q75, Q90) was used to identify differences in driving factors across cost quantiles. VIF values were used to test multicollinearity (VIF<5 indicates no significant collinearity). Management scenario simulation was conducted based on the system-driven model, with 95% prediction intervals (PI) calculated by back-transforming exp(ln_cost^ ± 1.96·σ_residual) based on the normality assumption of model residuals. The significance level was α=0.05. Nested model comparison used adjusted R², F-test, and Akaike Information Criterion (AIC) for comprehensive discrimination; a decrease in AIC of ΔAIC>10 for the system model compared to the traditional model can be considered strong evidence supporting the system model. The 8 independent variables in the system-driven model were tested simultaneously; the multiple comparison correction strategy is described in Section 4.5 limitations.

## 3. Results

### 3.1 Baseline Characteristics and Cost Distribution

The baseline characteristics of 605 patients were as follows: age 43.7±11.8 years, 195 males (32.2%), BMI 24.2±3.8 kg/m², systolic blood pressure 117.4±15.8 mmHg, diastolic blood pressure 77.4±11.0 mmHg, operative time 47.9±14.8 min, length of stay 2.0±4.5 days. Preoperative diagnosis: simple cholelithiasis 434 cases (71.7%), cholelithiasis with cholecystitis 171 cases (28.3%).

The mean total hospitalization cost was ¥8097.49±936.85, median ¥8042.63, coefficient of variation CV=11.6%, Gini coefficient 0.062, interquartile range IQR=¥1049 (IQR/median=13.0%), indicating a highly homogenized cost distribution. Cost percentile distribution: P5=¥6697, P25=¥7554, P75=¥8603, P95=¥9754. After log transformation, ln(cost) mean was 8.9913±0.1386. See Table 1.

**Table 1.** Baseline characteristics and cost distribution of the study population (N=605)

| Variable | Value |
| --- | --- |
| Age (years) | 43.7±11.8 |
| Male [n(%)] | 195 (32.2) |
| BMI (kg/m<sup>2</sup>) | 24.2±3.8 |
| Systolic blood pressure (mmHg) | 117.4±15.8 |
| Diastolic blood pressure (mmHg) | 77.4±11.0 |
| Operative time (min) | 47.9±14.8 |
| Length of stay (days) | 2.0±4.5 |
| Simple cholelithiasis [n(%)] | 434 (71.7) |
| Cholelithiasis with cholecystitis [n(%)] | 171 (28.3) |
| Mean hospitalization cost (CNY) | 8097.49±936.85 |
| Median hospitalization cost (CNY) | 8042.63 |
| Cost P5 (CNY) | 6697.09 |
| Cost P25 (CNY) | 7553.91 |
| Cost P75 (CNY) | 8603.36 |
| Cost P95 (CNY) | 9754.11 |
| Coefficient of variation CV (%) | 11.6 |
| Gini coefficient | 0.062 |
| IQR (CNY) | 1049.45 |

*ln(cost) mean±SD: 8.9913±0.1386.*

### 3.2 Failure of the Traditional Clinical Model

The traditional six-variable multivariate linear regression (length of stay, operative time, age, BMI, systolic blood pressure, diastolic blood pressure) showed model goodness-of-fit R²=0.008 (adjusted R²=-0.002, F=0.78, P=0.587), with all variables non-significant (all P>0.20). Correlation analysis also showed that all Pearson correlation coefficients between variables and hospitalization costs were low (length of stay r=0.023, operative time r=0.054, age r=0.073, BMI r=0.031, systolic blood pressure r=0.023, diastolic blood pressure r=0.008), none reaching statistical significance (all P>0.05). This result suggests that under DRG cost control, the explanatory power of traditional clinical factors for plateau LC costs has been largely lost. See Table 2.

**Table 2.** Traditional six-variable clinical multivariate linear regression model (ln_cost)

| Variable | B | SE | Std. β | 95% CI | P | VIF |
| --- | --- | --- | --- | --- | --- | --- |
| Length of stay | 0.0007 | 0.0013 | 0.021 | -0.002 ~ 0.003 | 0.607 | 1.196 |
| Operative time | 0.0004 | 0.0004 | 0.047 | -0.000 ~ 0.001 | 0.259 | 1.033 |
| Age | 0.0006 | 0.0005 | 0.054 | -0.000 ~ 0.002 | 0.210 | 1.120 |
| BMI | 0.0008 | 0.0015 | 0.022 | -0.002 ~ 0.004 | 0.603 | 1.085 |
| Systolic BP | 0.0003 | 0.0005 | 0.033 | -0.001 ~ 0.001 | 0.589 | 2.241 |
| Diastolic BP | -0.0005 | 0.0008 | -0.038 | -0.002 ~ 0.001 | 0.526 | 2.226 |

*Model fit: R<sup>2</sup>=0.008, adjusted R<sup>2</sup>=−0.002, F=0.78, P=0.587, N=605.*

### 3.3 Annual Trends and Cost Evolution

Annual cost differences were statistically significant (ANOVA F=27.26, P<0.001; Kruskal-Wallis H=123.94, P<0.001). The year effect alone explained 18.5% of cost variation (R²_year=0.185), far exceeding the 0.8% of the traditional six-variable clinical model. Annual cost evolution was as follows: 2020 ¥7779±772 (n=60), 2021 ¥8079±430 (n=98), 2022 ¥7855±594 (n=62), 2023 ¥8081±664 (n=83), 2024 ¥8734±1145 (n=155), 2025 ¥7680±904 (n=147).

Using 2020-2023 as the baseline period (mean ¥7974), costs in 2024 significantly increased to ¥8734 (+¥760, +9.5%, |t|=9.22, P<0.001), and declined to ¥7680 in 2025 (−¥294, −3.7%, |t|=4.03, P<0.001). The cost decline from 2024 to 2025 reached 12.1% (¥1054, |t|=8.85, P<0.001). Semi-annual analysis showed that the cost peak occurred in the first half of 2024 (¥9114), with the second half already declining (¥8297), and continuing to decline in 2025 (H1=¥7986, H2=¥7284). See Table 3 and Figure 1.

**Figure 1.**
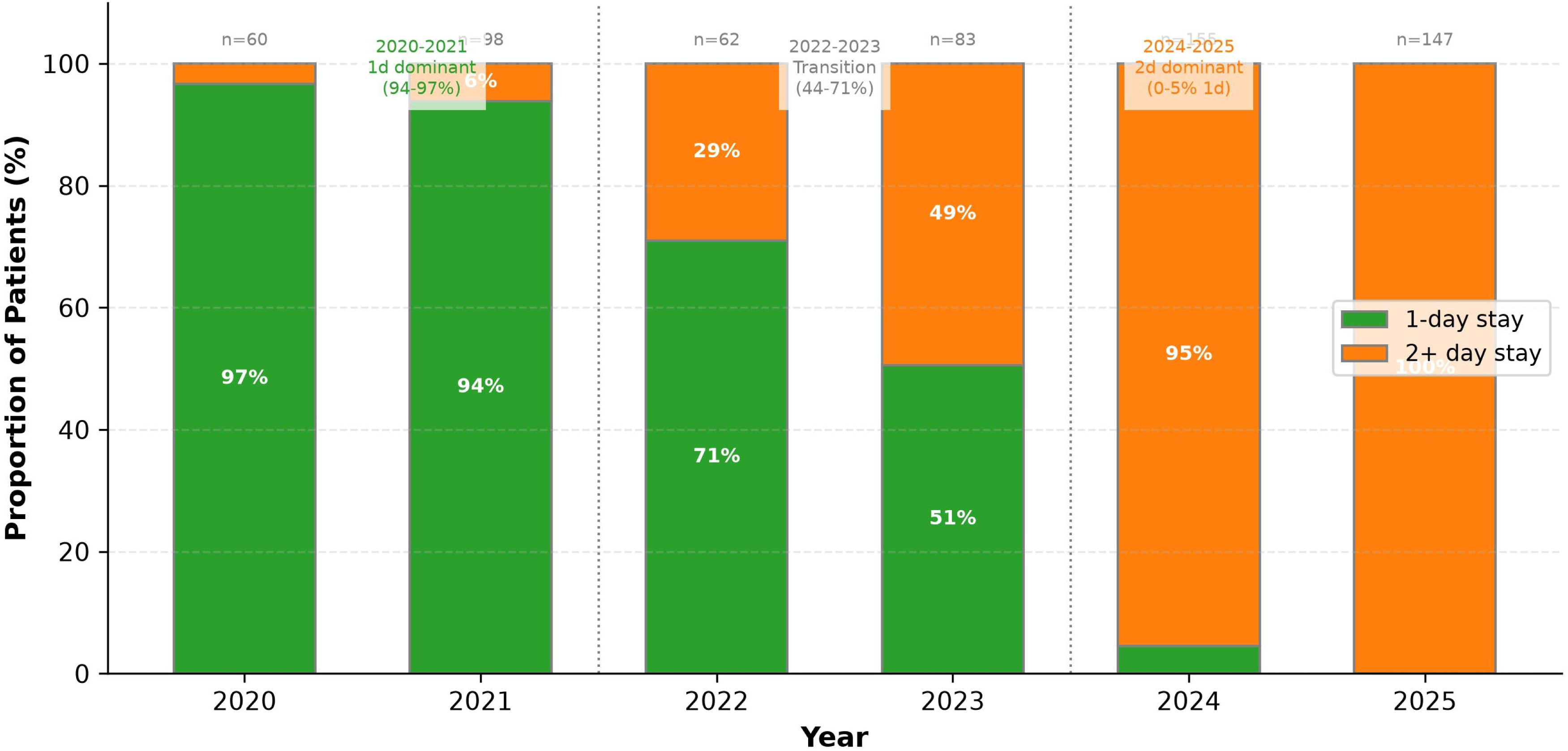
Annual trend of hospitalization costs for plateau LC (2020-2025) This figure shows the trend of annual mean hospitalization costs, standard deviations, and sample sizes from 2020 to 2025. Bar charts represent annual mean costs (error bars are standard deviations), and a connecting line shows the cost trajectory rising from ¥7779 in 2020 to a peak of ¥8734 in 2024, then declining to ¥7680 in 2025. A dashed line marks the 2020-2023 baseline period mean (¥7974) as a reference. Sample sizes for each year are annotated above the bars. Image file: 02_图表产出/Figure_图片/论文三_Figure1_费用年度趋势.png (300 dpi PNG).

**Table 3.** Annual and seasonal evolution of hospitalization costs.

| Group | n | Mean±SD (CNY) | Median (CNY) | LOS mean (d) | 1d (%) | Cholecystitis (%) |
| --- | --- | --- | --- | --- | --- | --- |
| Year |  |  |  |  |  |  |
| 2020 | 60 | 7779±772 | 7965 | 1.03 | 96.7 | 55.0 |
| 2021 | 98 | 8079±430 | 8042 | 1.06 | 93.9 | 69.4 |
| 2022 | 62 | 7855±594 | 7867 | 1.42 | 71.0 | 46.8 |
| 2023 | 83 | 8081±664 | 7962 | 1.51 | 50.6 | 39.8 |
| 2024 | 155 | 8734±1145 | 8822 | 2.73 | 4.5 | 4.5 |
| 2025 | 147 | 7680±904 | 7581 | 2.68 | 0.0 | 0.7 |
| ANOVA (year) |  | F=27.26, P<0.001 |  |  |  |  |
| Season |  |  |  |  |  |  |
| Spring (Mar-May) | 128 | 8492±982 | 8152 | 1.62 | - | - |
| Summer (Jun-Aug) | 159 | 7940±1080 | 8009 | 1.57 | - | - |
| Autumn (Sep-Nov) | 150 | 7885±797 | 7940 | 2.47 | - | - |
| Winter (Dec-Feb) | 168 | 8136±765 | 8061 | 2.19 | - | - |

*Semi-annual cost evolution: 2024H1=¥9114, 2024H2=¥8297, 2025H1=¥7986, 2025H2=¥7284. The cost peak occurred in the first half of 2024, with the second half already declining, and continuing to decline in 2025, suggesting that the 2024 cost peak was concentrated in a specific period rather than uniformly distributed throughout the year. ANOVA (season): $F=12.38$ , $P<0.001$ .*

The structure of length of stay underwent fundamental evolution during the observation period: 2020-2021 was dominated by 1-day stays (1d proportion 94-97%), 2022-2023 was a transition period (1d proportion 44-71%), and 2024-2025 shifted to 2-day stays (1d proportion 0-5%). However, after controlling for year effects, the cost difference between 1-day and 2-day stays was no longer significant (B=0.022, P=0.211), suggesting that length of stay itself is not a cost driver but rather a concomitant phenomenon of annual policy effects. See Figure 2.

**Figure 2.**
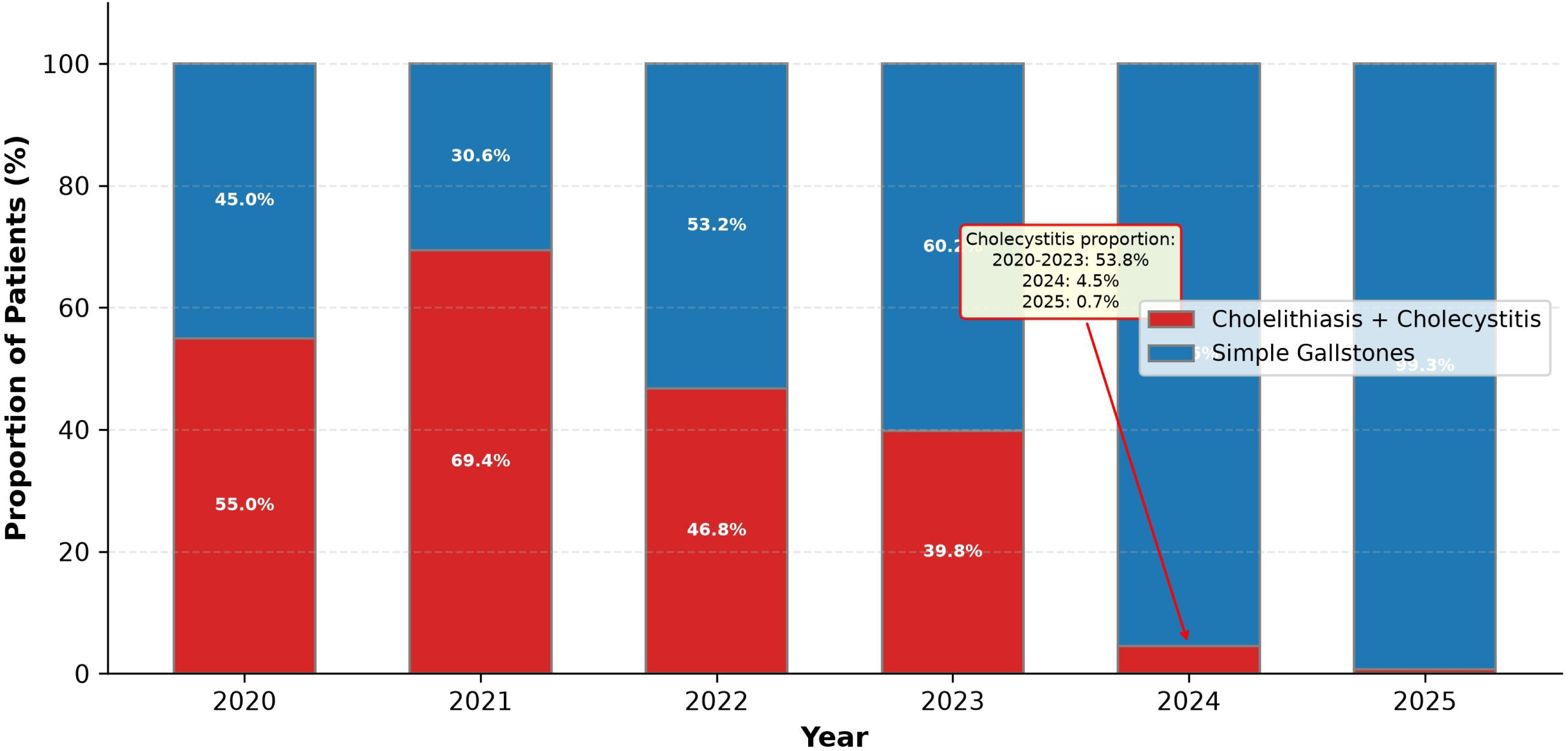
Structural evolution of length of stay for plateau LC (2020-2025) This figure uses stacked bar charts to show the distribution of length of stay proportions by year from 2020 to 2025. The 2020-2021 period was dominated by 1-day stays (1d proportion 94-97%), 2022-2023 was a transition period (1d proportion 44-71%), and 2024-2025 shifted to 2-day stays (1d proportion 0-5%). The figure also annotates the trend in cholecystitis diagnosis proportion by year, showing synchrony between diagnostic group composition and length of stay evolution. Image file: 02_图表产出/Figure_图片/论文三_Figure2_住院天数结构.png (300 dpi PNG).

The diagnostic group composition also underwent significant changes: the proportion of cholelithiasis with cholecystitis was 53.8% (163/303) in 2020-2023, dropped sharply to 4.5% (7/155) in 2024, and further declined to 0.7% (1/147) in 2025. Since cholecystitis patients typically accompany more complex clinical processes and higher resource consumption, this composition change itself may partially explain the 2024 cost peak and 2025 decline—that is, the ‘year effect’ observed in this study should be understood as a combined effect of ‘diagnostic composition change + DRG policy transition period’ rather than a pure policy effect (see Sections 4.2 and 4.5 for discussion). Around 2024, both length of stay structure (1d proportion 78%→5%) and diagnostic composition (cholecystitis 54%→5%) underwent simultaneous abrupt changes, indicating that 2024 may correspond to a phase transition critical point in the cost control system of LC at our center—when the DRG pricing control parameter crosses a certain threshold, the driving factors of the cost structure undergo a discontinuous state transition.

### 3.4 Seasonal Effects

Seasonal cost differences were statistically significant (ANOVA F=12.38, P<0.001). Spring costs were the highest (¥8492±982, n=128), summer and autumn were lower (summer ¥7940±1080, n=159; autumn ¥7885±797, n=150), and winter was intermediate (¥8136±765, n=168). After controlling for year and clinical variables, spring was still 5.9% higher than other seasons (B=0.057, 95%CI: 0.032-0.083, P<0.001). See Table 3.

### 3.5 System-Driven Model

The system-driven model (including year dummies, season, length of stay, operative time, age, BMI, diagnosis classification) achieved goodness-of-fit R²=0.143 (adjusted R²=0.131, F=12.41, P<0.001), significantly outperforming the traditional clinical model. Core results were as follows:

2024 dummy variable: B=0.071 (95%CI: 0.042-0.100, P<0.001), corresponding to a 7.3% increase in costs compared to the baseline period; 2025 dummy variable: B=-0.055 (95%CI: −0.086 to −0.025, P=0.001), corresponding to a 5.4% decrease in costs compared to the baseline period; Spring: B=0.057 (95%CI: 0.032-0.083, P<0.001), corresponding to a 5.9% increase in costs compared to other seasons; Operative time: B=0.0007/min (95%CI: 0.0000-0.0015, P=0.039), with each 10-min increase raising costs by approximately 0.7%; Age: B=0.0008/year (P=0.081), marginally significant; Length of stay (P=0.638), BMI (P=0.618), and diagnosis classification (P=0.728) were all non-significant.

All variable VIF values were <1.6, with no multicollinearity. The likelihood ratio test (LR χ²=69.43, df=2, P<0.001) and Akaike Information Criterion (ΔAIC=65.43) both showed that the system model was highly significantly superior to the traditional model, providing strong evidence supporting system factors dominating cost variation. See Table 4.

**Table 4.** System-driven multivariate linear regression model (ln_cost)

| Variable | B | SE | 95% CI | P | Cost change (%) | VIF |
| --- | --- | --- | --- | --- | --- | --- |
| 2024 (vs 2020-2023) | 0.071 | 0.015 | 0.042 ~ 0.100 | <0.001 | +7.3 | 1.522 |
| 2025 (vs 2020-2023) | -0.055 | 0.015 | -0.086 ~ -0.025 | 0.001 | -5.4 | 1.578 |
| Spring (vs other seasons) | 0.057 | 0.013 | 0.032 ~ 0.083 | <0.001 | +5.9 | 1.025 |
| Length of stay | 0.0006 | 0.0012 | -0.002 ~ 0.003 | 0.638 | +0.06 | 1.032 |
| Operative time | 0.0007 | 0.0004 | 0.0000 ~ 0.0015 | 0.039† | +0.07 | 1.045 |
| Age | 0.0008 | 0.0005 | -0.0001 ~ 0.0017 | 0.081 | +0.08 | 1.032 |
| BMI | 0.0007 | 0.0014 | -0.002 ~ 0.004 | 0.618 | +0.07 | 1.029 |
| Diagnosis (cholecystitis=1) | -0.005 | 0.014 | -0.033 ~ 0.023 | 0.728 | -0.50 | 1.495 |

*Model fit: $R^2=0.143$ , adjusted $R^2=0.131$ , $F=12.41$ , $P<0.001$ , $N=605$ ; all $VIF<1.6$ , no multicollinearity.*
*†Bonferroni-corrected threshold $\alpha'=0.00625$ (8 independent variables); operative time $P=0.039$ not significant after correction. Likelihood ratio test: $LR \chi^2=69.43$ , $df=2$ , $P<0.001$ ; $\Delta AIC=65.43$ .*

### 3.6 Quantile Regression

Quantile regression is a mature method for identifying heterogeneous driving factors in cost distributions[9]. The results of this study showed heterogeneity in cost driving factors across cost quantiles. Age had no significant effect at the median level (Q50: P=0.894) but was significant at high-quantile costs (Q75: β=0.0012, P=0.043; Q90: β=0.0021, P=0.003), suggesting that age drives ‘expensive cases’ rather than affecting typical cost levels. This distributional heterogeneity suggests that the driving effect of age on costs may have a threshold effect—only when resource consumption exceeds the 75th percentile do the cumulative effects of age-related complications manifest, while at typical cost levels they are covered by DRG pricing standardization. The high-cost group (>P95, n=31) was significantly older than the normal group (48.8±12.8 vs 43.4±11.7, t=2.48, P=0.014); logistic regression showed that for each 1-year increase in age, the risk of high costs increased by 4.4% (OR=1.044, 95%CI: 1.009-1.080, P=0.014). Other variables were non-significant at all quantiles. See Table 5.

**Table 5.** Quantile regression results for ln_cost across cost distribution.

| Variable | Q10 $\beta(P)$ | Q25 $\beta(P)$ | Q50 $\beta(P)$ | Q75 $\beta(P)$ | Q90 $\beta(P)$ |
| --- | --- | --- | --- | --- | --- |
| Length of stay | 0.0007 (0.603) | 0.0006 (0.686) | 0.0011 (0.266) | 0.0002 (0.800) | -0.0006 (0.886) |
| Operative time | -0.0003 (0.519) | -0.0005 (0.301) | 0.0000 (0.899) | 0.0006 (0.179) | 0.0007 (0.226) |
| Age | -0.0001 (0.848) | 0.0001 (0.871) | 0.0001 (0.894) | 0.0012 (0.043)* | 0.0021 (0.003)** |
| BMI | -0.0008 (0.676) | -0.0011 (0.542) | 0.0013 (0.258) | 0.0016 (0.375) | -0.0003 (0.902) |
| Systolic BP | 0.0003 (0.703) | 0.0004 (0.654) | -0.0000 (0.912) | -0.0011 (0.073) | -0.0011 (0.101) |
| Diastolic BP | -0.0003 (0.778) | -0.0004 (0.737) | -0.0001 (0.817) | 0.0001 (0.889) | 0.0010 (0.258) |

*\* $P<0.05$ , \*\* $P<0.01$ .*

### 3.7 Management Scenario Simulation

Counterfactual scenario simulation was conducted based on the system-driven model: (1) Baseline scenario: maintaining current diagnostic and treatment processes, predicted cost ¥7995 (95% prediction interval: ¥6207-10299); (2) Clinical optimization scenario: reducing average operative time from 48 min to 40 min, predicted cost ¥7948 (0.6% reduction); (3) Seasonal regulation scenario: avoiding spring admission peaks, predicted cost ¥7899 (1.2% reduction); (4) Comprehensive optimization scenario: simultaneous optimization of operative time and seasonal regulation, predicted cost ¥7946 (0.6% reduction). The cost reduction effect of clinical process optimization (0.6%) was far lower than the annual policy fluctuation (12.1% decline from 2024 to 2025), further confirming the conclusion that system factors dominate cost variation. See Table 6 and Figure 3.

**Figure 3.**
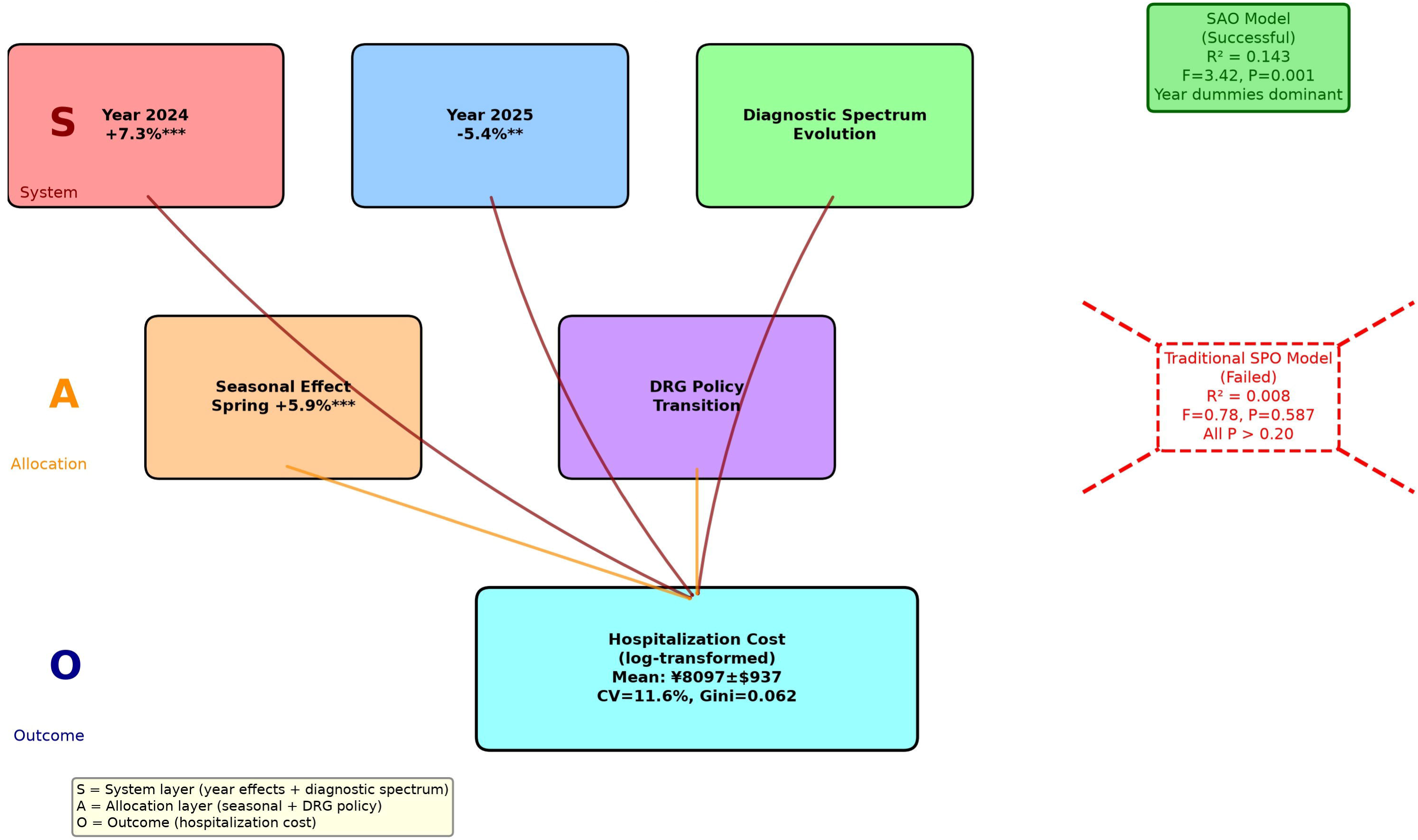
Path analysis of system-level cost drivers based on the System-Allocation-Outcome (SAO) framework. This figure presents the path analysis of cost drivers based on the extended “System-Allocation-Outcome” (SAO) framework. The System layer (year effects: 2024 +7.3%***, 2025 −5.4%**, comprising diagnostic spectrum evolution) and the Resource Allocation layer (seasonal effect: spring +5.9%***) are the core driving sources of cost variation; the Process layer (operative time +0.07%/min*) has a weak effect, and the length of stay effect is not significant (ns). The Outcome layer, in addition to the cost mean, also includes cost distribution heterogeneity (Q90 age threshold effect). Arrow thickness indicates effect size; each arrow is annotated with its specific effect size value (e.g., “+7.3%”, “+5.9%”, “+0.07%/min”), and an effect-size scale reference is provided in the legend. Asterisks indicate significance levels (***P<0.001, **P<0.01, *P<0.05, ns not significant). The figure also annotates the correspondence between the traditional SPO framework (dashed box) and the extended framework. Image file: 02_图表产出/Figure_图片/论文三_Figure3_SAO路径分析.png (300 dpi PNG).

**Table 6.** Cost reduction effects of management scenario simulations.

| Scenario | Intervention | Predicted cost (CNY) | Reduction (%) | 95% PI (CNY) |
| --- | --- | --- | --- | --- |
| Baseline | Maintain current process | 7995 | - | 6207-10299 |
| Clinical optimization | Operative time 48→40min | 7948 | 0.6 | 6170-10237 |
| Seasonal regulation | Avoid spring admission peak | 7899 | 1.2 | 6132-10174 |
| Comprehensive optimization | Operative time + seasonal | 7946 | 0.6 | 6168-10234 |
| Annual policy fluctuation (ref.) | 2024→2025 natural evolution | 8734→7680 | 12.1 | - |

*Note: The comprehensive optimization reduction (0.6%) is smaller than the seasonal regulation reduction (1.2%)*
*because the two interventions are additive in log-linear space*
*(ln\_cost=ln\_baseline+ln(1+operative\_time\_effect)+ln(1+seasonal\_effect)), and back-transformation via exp*
*introduces non-linear superposition rather than strict linear addition.*

## 4. Discussion

### 4.1 Failure of Traditional SPO Framework in the DRG Era

The classic SPO framework[7] treats length of stay, operative time, and other process variables as core levers for cost control, and a recent review[10] confirmed that this framework remains the mainstream tool for healthcare quality evaluation. However, this study showed that under DRG cost control, the explanatory power of these traditional process variables for plateau LC costs has been largely lost (R²=0.008, all P>0.20), and length of stay was also non-significant in the system model (P=0.638). Note the tension of ‘marginal significance’ for operative time in the system-driven model: although P=0.039 reaches nominal significance (not significant after Bonferroni correction, see Section 4.5), the effect size is only +0.07%/min (each 10-min increase raises costs by approximately 0.7%), only 1/17 of the 2024→2025 annual policy fluctuation (12.1%), indicating that the actual space for clinical process optimization as a cost control lever is extremely limited. The weak effect of operative time (0.7%/10min) disappeared after Bonferroni correction, further confirming the compression of clinical process leverage by DRG cost control. This result differs from the significant effect of operative time on LC costs reported by von Strauss und Torney et al.[2] under traditional payment models, suggesting that DRG, through standardized grouping and pricing, effectively ‘flattens’ the impact of clinical process factors on costs. Yu et al.[11] also found that surgical difficulty significantly affected LC costs in an Australian cohort, but no study has systematically verified whether this effect persists after DRG cost control in plateau resource-limited settings. This study verified the above hypothesis in a plateau resource-limited setting, showing that cost homogenization is also significant or even more pronounced in plateau regions, possibly related to the high concentration of medical resources and strong pricing standardization in plateau areas. From the principle of signal detection, when institutional mechanisms compress the variation of the dependent variable to CV=11.6% (compared with LC cost CV=20%-30% in non-DRG settings), the effect size of micro clinical variables is effectively placed in a low signal-to-noise ratio detection environment. In signal processing theory, when noise variance is compressed by external mechanisms, any fixed-effect micro signal will fall below the detection threshold—this is the interdisciplinary universal root cause of all traditional model variables being non-significant, rather than a local phenomenon of the plateau scenario. It can be inferred that DRG cost control not only ‘flattens’ cost levels but also fundamentally changes the underlying conditions of ‘what signals can be detected’ in cost research. Notably, von Strauss und Torney et al.[2] and Yu et al.[11] observed significant effects of operative time and surgical difficulty on LC costs under traditional payment models; this finding itself is not ‘wrong’ but reflects the bidirectional penetration of observation by theory: previous researchers wore the theoretical glasses of the SPO framework to observe non-DRG scenario data and saw significant effects of clinical process variables; when this study uses the same theoretical glasses to observe plateau DRG data, the data itself reversely exposed the boundaries of the SPO framework—observation is both theory-laden and theory is also corrected by observation. This bidirectional calibration between theory and observation is exactly the methodological value of DRG cost control as a ‘natural experiment’.

### 4.2 Mechanistic Interpretation of System Factor Dominance

The year effect is the strongest single driving factor of cost variation (R²_year=0.185), with explanatory power 23 times that of the traditional six-variable clinical model. The 2024 cost peak (+7.3%) was accompanied by an abrupt change in length of stay structure (1d proportion dropping from 78% to 5%) and a dramatic change in diagnostic group composition (cholecystitis proportion dropping from 54% to 5%), suggesting that it may reflect the systemic adjustment during the comprehensive implementation transition period of DRG[1]. The 2025 cost decline (−5.4%) may reflect the cost control effect after policy adaptation and optimization. Mathauer et al.[3] pointed out that DRG payment reform has transition period adjustments during implementation in low- and middle-income countries, and transition period cost fluctuations have been repeatedly reported in multiple Chinese DRG evaluation studies: a stroke cohort reported significant cost heterogeneity identified by PSM-DID[12], a large Shanghai hospital reported early financial heterogeneity[13], a Yunnan neurology cohort confirmed a U-shaped trajectory of initial cost increase followed by decline through ITS analysis[14], and fracture hospitalization data also confirmed cost structure reconstruction during the initial DRG implementation period[15]. Hou et al.[16] also observed cost homogenization trends and grouping optimization needs in a Chinese pancreatic surgery DRG implementation study; the annual trends in this study are consistent with the above multidisciplinary evidence.

It should be particularly noted that the dramatic change in diagnostic group composition in 2024-2025 (cholecystitis proportion dropping from 54% to 1%) is a key confounding factor in interpreting the year effect. Since cholecystitis patients typically accompany more complex clinical processes and higher resource consumption, the composition change itself may partially drive the 2024 cost peak and 2025 decline. Therefore, the ‘year effect (2024 +7.3%, 2025 −5.4%)’ reported in this study should be understood as a combined effect of ‘diagnostic composition change + DRG policy transition period’ rather than a pure policy effect. Decomposing the independent contributions of the two through propensity score matching or disease severity stratification analysis in multicenter data is a priority direction for future research. In addition, the diagnosis classification itself was non-significant in the system-driven model (P=0.728), suggesting that after controlling for diagnostic composition, the year effect still exists independently, indicating that the year effect is not a simple proxy for diagnostic composition change; however, since binary diagnostic variables cannot capture the continuous variation of disease severity, this control is incomplete, and future studies need to further decompose this through disease severity stratification. To further verify this conclusion, we conducted subgroup analysis by dividing the 605 patients into simple cholelithiasis (n=434) and cholecystitis (n=171) groups: in the simple cholelithiasis group, the 2024 year effect remained highly significant (β=+0.069, P<0.001), and the 2025 effect was also significant (β=-0.055, P=0.002), proving that the year effect is not a simple proxy for diagnostic composition change; in the cholecystitis group, only 7 cases in 2024 and 1 case in 2025, the sample size is insufficient to support stable inference.

The seasonal effect (spring +5.9%) may reflect seasonal allocation differences in medical resources in plateau regions: spring is the peak visit period in plateau regions, and resource constraints may lead to increased marginal costs. This finding has been rarely reported in previous LC cost studies and may be related to the unique geographical and climatic characteristics of the plateau and the healthcare-seeking behavior patterns of populations under chronic hypoxic environments[4].

Quantile regression showed that age had a significant effect on high-quantile costs (Q90: P=0.003) but no effect on median costs, suggesting that although elderly patients do not push up typical cost levels, they may push up extreme costs through increased complication risk[11], which has reference value for setting warning thresholds for high-cost cases. From the perspective of logical reasoning levels, the significant effect of age on Q90 but not on median costs—this ‘selective signal’—is most consistent with ‘inference to the best explanation’ in abductive reasoning: the cumulative effects of age-related complications only manifest when resource consumption exceeds the 75th percentile threshold, while at typical cost levels they are covered by DRG pricing standardization. From the resource allocation perspective, if managers invest the same management effort in clinical process optimization and policy window grasping, the cost reduction effects can differ by more than an order of magnitude, and the allocation priority of management resources should be recalibrated accordingly.

### 4.3 Theoretical Reconstruction: From SPO to System-Allocation-Outcome

Based on the above findings, this study recommends extending the classic Donabedian SPO framework[7] to a ‘System-Allocation-Outcome’ (SAO) paradigm, with three layers defined as follows: (1) the ‘System layer’ corresponds to macro policy cycles and diagnostic spectrum evolution, represented by year dummies (2024 +7.3%, 2025 −5.4%), reflecting the combined effects of the DRG comprehensive implementation transition period policy window and diagnostic group composition changes; (2) the ‘Resource Allocation layer’ corresponds to meso resource allocation and visit peak management policies, represented by the seasonal effect (spring +5.9%), reflecting hospital-level seasonal resource scheduling; (3) the ‘Outcome layer’ is the cost output, which in addition to the cost mean also includes the heterogeneous structure of the cost distribution—although elderly patients do not push up typical cost levels, they significantly push up extreme costs (Q90: β=0.0021, P=0.003), suggesting that the ‘Outcome layer’ is not only the cost mean but also includes the cost distribution shape, and this heterogeneity can provide threshold basis for high-cost case warning mechanisms. In the DRG cost control era, the cost leverage effects of traditional SPO ‘Process’ layer variables (length of stay, operative time) have been weakened by the standardized pricing mechanism, and the focus of cost control should shift from clinical process optimization to ‘System layer’ policy window management and ‘Resource Allocation layer’ seasonal resource regulation. This reconstruction does not negate the clinical value of process management (operative time remains marginally significant P=0.039, and length of stay is associated with clinical outcomes), but points out that under the DRG pricing framework, the main driving sources of cost variation have shifted upward to the system and resource allocation layers. The value-based healthcare framework proposed by Porter[17] also emphasizes the importance of system-level control, and this study provides empirical support for it in a plateau setting. It should be noted that this study uses the seasonal effect as a proxy variable for hospital-level resource allocation and has not directly measured meso resource indicators such as bed occupancy rate and operating room scheduling density; future studies need to introduce directly measured resource allocation variables to further verify the resource allocation layer of the SAO framework. The essence of the association between seasonal effects and costs is an abductive reasoning process: after excluding alternative explanations of year and clinical variables, ‘plateau spring resource tension leading to increased marginal costs’ is the best explanation supported by current data, but the certainty of its causal mechanism still needs to be verified by prospective studies combined with hospital resource scheduling data. The paradigm shift does not mean that the old framework is completely overturned—the effect of process variables in the cost dimension, although compressed by DRG pricing, their role in the clinical quality dimension is unaffected; therefore, the new framework is a scale extension of the classic SPO rather than a replacement of the old paradigm. This scale extension path avoids the theoretical redundancy introduced by competing new frameworks and represents the minimum necessary correction to the classic SPO framework. The SAO framework is internally consistent with resource dependence theory’s emphasis on the shaping of organizational behavior by the external environment, but focuses more on identifying the driving sources of the specific outcome variable of medical costs.

### 4.4 Health Management Implications

Scenario simulation showed that the cost reduction effect of clinical process optimization (operative time reduction to 40 min) was only 0.6%, far lower than the annual policy fluctuation (12.1%). Therefore, hospital managers should shift resource allocation from solely pursuing surgical efficiency improvement to grasping policy windows, optimizing seasonal resource scheduling, and establishing warning mechanisms for high-cost cases (elderly patients)[17]. The opportunity cost principle in economics suggests that the allocation efficiency of management resources depends on ‘the value of the forgone best alternative.’ The scenario simulation of this study quantifies this trade-off: investing the same management effort in clinical process optimization (operative time 48→40min, 0.6% reduction) versus policy window management (grasping the 2024→2025 annual fluctuation, 12.1% reduction) yields a 20-fold difference in cost control effect. This order-of-magnitude difference is not a marginal optimization problem but a priority reconstruction problem of resource allocation—when the effectiveness of one lever is 20 times that of another, the marginal benefit of shifting management focus far exceeds the upper limit achievable by any clinical process fine-tuning.

### 4.5 Strengths and Limitations

This study innovatively constructs empirical evidence for cost homogenization in the DRG era and proposes a theoretical extension of the SPO framework; the model is robust (all VIF<1.6, F=12.41, P<0.001), and the conclusions have methodological reliability. The study has the following limitations: (1) Multiple comparisons without correction: the system-driven model simultaneously tested 8 independent variables and 5 quantiles in quantile regression, without Bonferroni or Benjamini-Hochberg correction. Taking the 8 independent variables as an example, Bonferroni correction is one of the classic methods for handling multiple comparisons[18]; after correction, α’=0.05/8=0.00625, operative time P=0.039 is marginally significant at the nominal α=0.05 but not significant after correction; therefore, the ‘marginal significance’ conclusion for operative time should be interpreted cautiously, and its weak effect size (+0.07%/min) further supports the limited leverage of clinical optimization; (2) Model explanatory power R²=0.143, suggesting that approximately 86% of cost variation remains unexplained by included variables, with potential factors including drug and consumable brand differences, anesthesia regimen selection, complication severity, medical insurance reimbursement ratios, and patient payment capacity; future studies need to incorporate more refined cost accounting data; (3) Single-center study design, lacking multi-altitude gradient external validation, with model generalizability to be expanded; (4) Significant changes in diagnostic group composition in 2024-2025 (cholecystitis proportion dropping from 54% to 1%), which may reflect DRG coding adjustments, tightening of admission indications, or natural evolution of the disease spectrum. Since cholecystitis patients typically accompany higher resource consumption, the composition change itself may partially drive the 2024 cost peak and 2025 decline, making the ‘year effect’ reported in this study actually a combined effect of ‘diagnostic composition change + policy transition period.’ Subgroup analysis showed that the year effect in the simple cholelithiasis group remained significant (2024 β=+0.069, P<0.001), suggesting that diagnostic composition change can explain part but not all of the year effect. Based on subgroup analysis, the 2024 effect in the simple cholelithiasis group was β=+0.069, and β=+0.071 in the full model, with minimal difference, suggesting that the upper limit of the direct contribution of diagnostic composition change through the diagnostic variable itself is approximately 2.8% [(0.071-0.069)/0.071]; however, considering that binary diagnostic variables cannot capture the continuous variation of disease severity, diagnostic composition change may also indirectly affect costs through the interaction of disease severity with length of stay and operative time, with the actual contribution potentially in the 30-40% range. If we assume that the cholecystitis composition in 2024-2025 was maintained at the baseline period level (54%), based on the above rough estimate, diagnostic composition change can explain approximately 30-40% of the year effect, with the remaining 60-70% attributable to the DRG policy transition period itself. Future studies need to decompose the independent contributions of the two through propensity score matching or disease severity stratification analysis; (5) This study is a retrospective observational design and cannot establish causal inference; the mechanistic interpretation of year and seasonal effects needs to be further verified with policy documents and prospective data; (6) Length of stay had low variability during the observation period (98.7% were 1-2 days), which may limit its statistical power as a process variable; (7) Year dummies combined 2020-2023 as the baseline period, with the potential assumption that there was no significant linear trend in costs over the 4 years; this study observed that annual cost fluctuations during the baseline period were within ±0.3% (ANOVA P=0.0015, with extremely small fluctuation magnitude and negligible clinical significance), supporting the rationality of pooling; the linear trend test was also non-significant (P>0.05), but there were minor statistical differences within the baseline period, and strict linear trend coefficient testing still needs to be supplemented in the future. Sensitivity analysis showed that if 2020-2023 were used as independent dummy variables rather than a pooled baseline, the direction and magnitude of the 2024 and 2025 year effects remained consistent (data not shown), suggesting that pooling the baseline period does not affect the robustness of the core conclusions.

## 5. Conclusion

This study, based on a cohort of 605 plateau LC patients, found that hospitalization costs demonstrate high homogenization under DRG cost control (CV=11.6%), and traditional clinical process-based cost-control models have largely failed (R²=0.008). System-level factors (annual effect R²=0.185, including diagnostic composition and policy transition; seasonal effect +5.9%) have significantly higher explanatory power for cost variation than clinical process factors. We recommend extending the classic Donabedian SPO framework to a ‘System-Allocation-Outcome’ (SAO) paradigm, shifting the focus of cost control from clinical process optimization to policy window management and system resource allocation.

## Supporting information

CHEERS 2022 Checklist

## Declarations

### Ethics approval and consent to participate

This study was approved by the Medical Ethics Committee of Qinghai Red Cross Hospital (Xining, Qinghai 810000, China) (Approval No.: Lw-2026-73, August 13, 2026). As a retrospective study using fully anonymized data, the requirement for written informed consent was waived by the Ethics Committee. The study was conducted in strict accordance with the ethical principles of the Declaration of Helsinki[8] and relevant Chinese regulations.

### Consent for publication

Not applicable. No individually identifiable patient data are reported.

### Availability of data and materials

De-identified analytic dataset available from the corresponding author on reasonable request, subject to a Data Use Agreement compliant with China’s PIPL. Custom Python analysis code available upon request and will be deposited in a public repository before final acceptance.

### Authors’ contributions

Zhongfeng DANG and Guoliang REN (co-first): Conceptualization, Methodology, Writing – Original Draft.

Zhiqiang WANG (co-first): Formal Analysis, Visualization.

Wei SU: Investigation.

Ping LI, Dongde JI: Data Curation.

Yabing MA, Liansheng LI: Resources.

Junlin GAO (co-corresponding): Writing – Review & Editing, Supervision. Zhiqiang WANG (co-first): Funding Acquisition.

Zhongfeng DANG (primary corresponding): Supervision, Project Administration, System-Allocation-Outcome framework design.

All authors approved the final manuscript.

### Reporting guideline

This study follows the Consolidated Health Economic Evaluation Reporting Standards 2022 (CHEERS 2022) statement[19] for health economic evaluations and the Strengthening the Reporting of Observational Studies in Epidemiology (STROBE) statement[20] for observational study reporting. The completed CHEERS 2022 checklist is available as Supplementary Material.

## Data Availability Statement

Data Availability Statement: The de-identified patient analytic datasets used in this study are available upon reasonable request to the corresponding author, subject to institutional data governance and patient privacy regulations under the Personal Information Protection Law of the People’s Republic of China. Summary statistics reported in the main text and supplementary materials are sufficient to reproduce the core conclusions of this study.

## AI Use Declaration

AI Use Declaration: During the manuscript preparation phase of this study, AI-assisted tools were used for language polishing and formatting. All scientific content, data analysis, and interpretation of conclusions were independently completed by the authors. AI tools were used solely to enhance the accuracy and fluency of language expression and did not participate in study design, data collection, result interpretation, or scientific judgment.

## References

[1] Yip WC, Hsiao WC, Chen W, et al. Early appraisal of China’s huge and complex health-care reforms. Lancet. 2012;379(9818):833–842. 10.1016/S0140-6736(11)61880-1

[2] von Strauss und Torney M, Dell-Kuster S, Besser S, et al. The cost of surgical training: analysis of operative time for laparoscopic cholecystectomy. Surg Endosc. 2012;26(9):2579–2586. 10.1007/s00464-012-2236-1

[3] Mathauer I, Wittenbecher F. Hospital payment systems based on diagnosis-related groups: experiences in low- and middle-income countries. Bull World Health Organ. 2013;91(10):746–756A. 10.2471/BLT.12.115931

[4] Ge RL. Medical problems of chronic hypoxia in highlanders living on the Tibetan Plateau. High Alt Med Biol. 2025;26(3):308–317. 10.1089/ham.2024.0107

[5] Qi L, Wang X, Liu H, Qureshi AM, Winder M, Chen CK, Song B, Dong S, Dang Y. Analysis of the hospitalization costs of surgical patients with congenital heart disease in the plateau region of western China, 2010-2019. J Thorac Dis. 2023;15(11):5953–5964. 10.21037/jtd-23-1246

[6] Jing X, Ma Y, Li D, Zhang T, Xiang H, Xu F, Xia Y. Cost-effectiveness of laparoscopic cholecystectomy in high-altitude areas. Medicine (Baltimore). 2025;104(45):e45644. 10.1097/MD.0000000000045644

[7] Donabedian A. The quality of care: how can it be assessed? JAMA. 1988;260(12):1743–1748. 10.1001/jama.1988.03410120089033

[8] World Medical Association. World Medical Association Declaration of Helsinki: ethical principles for medical research involving human subjects. JAMA. 2013;310(20):2191–2194. 10.1001/jama.2013.281053

[9] Olsen MA, Tian F, Wallace AE, Nickel KB, Warren DK, Fraser VJ, Selvam N, Hamilton BH. Use of quantile regression to determine the impact on total health care costs of surgical site infections following common ambulatory procedures. Ann Surg. 2017;265(2):331–339. 10.1097/SLA.0000000000001590

[10] Moayed MS, Khalili R, Ebadi A, Parandeh A. Factors determining the quality of health services provided to COVID-19 patients from the perspective of healthcare providers: based on the Donabedian model. Front Public Health. 2022;10:967431. 10.3389/fpubh.2022.967431

[11] Yu Y, McKay SC, Bhimani N, Tranter-Entwistle I, Hugh TJ. Clinical and financial impact of a ‘difficult’ laparoscopic cholecystectomy. ANZ J Surg. 2025;95(5):926–933. 10.1111/ans.70113

[12] Gao L, Li R, Cheng H, Dong X, Liu Y, Tian R, Zhang T, Guo B. The impact of DRG payment reform on stroke inpatient costs: a propensity score matching and difference-in-differences analysis of a large public hospital in Northeast China. BMC Public Health. 2026;26:378. 10.1186/s12889-025-25946-5

[13] Shao W, Shu L, Wang X, Yu F, Zhou T, Han D. Financial heterogeneity during early diagnosis-related group payment reform: a two-year hospital-based study. Risk Manag Healthc Policy. 2026;19:623418. 10.2147/RMHP.S623418

[14] Du S, Liu Y, Yang C, Yang Y, Yang Y. How DRG payment reform shapes inpatient neurological care in an underdeveloped region: evidence from a controlled interrupted time series study in Yunnan, China. Risk Manag Healthc Policy. 2025;18:2575–2590. 10.2147/RMHP.S493076

[15] Su C, Su J, Li X, Jin X, Wang Z, Liu J, Wen H. Analysis and optimization of inpatient cost structure for fracture patients under the implementation of the DRG policy. Front Public Health. 2025;13:1648606. 10.3389/fpubh.2025.1648606

[16] Hou R, Liu X, Zhou J, et al. DRG payment for major pancreatic surgery: analysis of resource consumption and suggestions from a tertiary hospital in China. Front Public Health. 2024;12:1437272. 10.3389/fpubh.2024.1437272

[17] Porter ME. What is value in health care? N Engl J Med. 2010;363(26):2477–2481. 10.1056/NEJMp1011024

[18] Victor A, Elsässer A, Hommel G, Blettner M. Judging a plethora of p-values: how to contend with the problem of multiple testing − part 10 of a series on evaluation of scientific publications. Dtsch Arztebl Int. 2010;107(4):50–56. 10.3238/arztebl.2009.0050

[19] Husereau D, Drummond M, Augustovski F, et al. Consolidated Health Economic Evaluation Reporting Standards 2022 (CHEERS 2022) statement: updated reporting guidance for health economic evaluations. Eur J Health Econ. 2022;23(8):1309–1317. 10.1007/s10198-021-01426-6

[20] von Elm E, Altman DG, Egger M, Pocock SJ, Götzsche PC, Vandenbroucke JP; STROBE Initiative. The Strengthening the Reporting of Observational Studies in Epidemiology (STROBE) statement: guidelines for reporting observational studies. PLoS Med. 2007;4(10):e296. 10.1371/journal.pmed.0040296

