## Supplementary material for "Cost Homogenization and System-Level Drivers in Plateau Laparoscopic Cholecystectomy: Failure and Reconstruction of Traditional Cost-Control Models in the DRG Era": CHEERS 2022 Checklist

### CHEERS 2022 Checklist — Paper 3

Reporting Guideline: CHEERS 2022 (Consolidated Health Economic Evaluation Reporting Standards 2022)

Checklist Version: CHEERS 2022 (consensus-based recommendation)

Last Updated: 2026-08-16

---

#### Checklist

| Item | Recommendation | Page/Section | Reported |
|---|---|---|---|
| **Title and abstract** | | | |
| 1 | Title identifies the study as an economic evaluation | Title (Cost Homogenization... Health Economics) | ✅ |
| 2 | Abstract provides a structured summary | Abstract (Background/Methods/Results) | ✅ |
| **Introduction** | | | |
| 3 | Provide statement of the background and context of the study | Introduction (DRG/DIP reform) | ✅ |
| 4 | State the research question, study perspective, and objectives | Introduction (Objectives) | ✅ |
| **Methods** | | | |
| 5 | Describe the population, intervention, comparator, and context | Methods (Population) | ✅ |
| 6 | State the time horizon and discount rate | Methods (Time Horizon: 2020–2025) | ✅ |
| 7 | Describe the chosen form of economic evaluation and analytical approach | Methods (Cross-sectional cost analysis) | ✅ |
| 8 | Describe the chosen perspective | Methods (Health system perspective) | ✅ |
| 9 | Describe the alternatives being compared | Methods (Traditional clinical model vs system-driven model) | ✅ |
| 10 | Describe the design and characteristics of the data sources | Methods (Single-center cohort) | ✅ |
| 11 | Describe selection of participants, exposures, and outcomes | Methods (Inclusion criteria) | ✅ |
| 12 | Describe identification and valuation of outcomes | Methods (Hospitalization cost outcomes) | ✅ |
| 13 | Describe measurement of | Methods (Cost measurement) | ✅ |

| | | | |
|---|---|---|---|
| | costs | | |
| 14 | Describe valuation of costs | Methods (CNY valuation) | ✅ |
| 15 | Describe analytical methods | Methods (Linear regression, quantile regression) | ✅ |
| 16 | Describe approaches to characterize uncertainty | Methods (95% CI, sensitivity analysis) | ✅ |
| 17 | Describe approaches to characterize heterogeneity | Methods (Subgroup analysis) | ✅ |
| **Results** | | | |
| 18 | Report the study sample | Results (Participants) | ✅ |
| 19 | Report summary results for each alternative | Results (Cost summary, CV, Gini) | ✅ |
| 20 | Report results from uncertainty analyses | Results (95% CI) | ✅ |
| 21 | Report results from heterogeneity analyses | Results (Year dummies) | ✅ |
| **Discussion** | | | |
| 22 | Summarize key findings | Discussion (Key Findings) | ✅ |
| 23 | Discuss limitations | Discussion (Limitations) | ✅ |
| 24 | Discuss generalizability | Discussion (Generalizability) | ✅ |
| 25 | Discuss implications for future research | Discussion (SAO paradigm) | ✅ |
| **Other information** | | | |
| 26 | State source of funding and role of the funder | Funding (YNZXKT2026009) | ✅ |
| 27 | State conflicts of interest | COI Statement | ✅ |

---

### Notes

- Health economic evaluation (cost analysis under DRG/DIP payment reform).
- Qinghai Red Cross Hospital, May 2020 – October 2025.
- Total participants: 605 plateau LC patients.
- Key findings: CV=11.6%, Gini=0.062 (high homogenization); traditional clinical model $R^2=0.008$; system-driven model $R^2=0.143$.
- Proposes SAO (System-Allocation-Outcome) paradigm to replace traditional SPO framework.
- Ethics Approval: LW-2026-71 (Qinghai Red Cross Hospital Medical Ethics Committee, 2026-08-15).

---

Checklist completed on 2026-08-16 by Zhongfeng DANG.
